# COVID-19: The unreasonable effectiveness of simple models

**DOI:** 10.1101/2020.05.26.20110957

**Authors:** Timoteo Carletti, Duccio Fanelli, Francesco Piazza

## Abstract

When the novel coronavirus disease SARS-CoV2 (COVID-19) was officially declared a pandemic by the WHO in March 2020, the scientific community had already braced up in the effort of making sense of the fast-growing wealth of data gathered by national authorities all over the world. However, despite the diversity of novel theoretical approaches and the comprehensiveness of many widely established models, the official figures that recount the course of the outbreak still sketch a largely elusive and intimidating picture. Here we show unambiguously that the dynamics of the COVID-19 outbreak belongs to the simple universality class of the SIR model and extensions thereof. Our analysis naturally leads us to establish that there exists a fundamental limitation to any theoretical approach, namely the unpredictable non-stationarity of the testing frames behind the reported figures. However, we show how such bias can be quantified self-consistently and employed to mine useful and accurate information from the data. In particular, we describe how the time evolution of the reporting rates controls the occurrence of the apparent epidemic peak, which typically follows the true one in countries that were not vigorous enough in their testing at the onset of the outbreak. The importance of testing early and resolutely appears as a natural corollary of our analysis, as countries that tested massively at the start clearly had their true peak earlier and less deaths overall.

## Introduction

In December 2019 the novel coronavirus disease SARS-CoV2 (COVID-19) emerged in Wuhan, China. Despite the drastic containment measures implemented by the Chinese government, the disease quickly spread all over the world, officially reaching pandemic level in March 2020.^1^ From the beginning of the outbreak, the international community braced up in the commendable effort to rationalise the massive load of data,^2,3^ painstakingly collected by local authorities throughout the world. However, epidemic spreading is a complex process, whose details depend to a large extent on the behaviour of the virus^4,5^ and of its current principal vectors (we, the humans). Additionally, recorded data necessarily suffer from many kinds of bias, so that much information is intrinsically missing in the data released officially. For example, the total number of reported infected bears the imprint of the growing size of the sampled population. This major source of non-stationarity should be properly gauged if one aims at explaining the observed trends and elaborate forecasts for the epidemics evolution. Similarly, the time series of recovered individuals that are released officially are heavily unreliable, due to the uneven ability of different countries to trace asymptomatic or mildly symptomatic patients.

In this letter, we demonstrate that overlooking these effects inevitably leads to fundamentally wrong predictions, irrespective of whether mean-field, compartment-based models^6,7^ or more sophisticate individual-based, stochastic spatial epidemic models^8,9^ are employed. More precisely, it is important to realise that the populations of infected reported by national agencies do not provide a reliable picture to correctly identify the epidemics peaks. The primary cause resides in the marked *non-stationary* character of the testing activity. However, while sources of bias due to differences in sampling and reporting policies among different countries are generally acknowledged, notably by studies applying Bayesian inference ^10,11^ and causal models,^12^ surprisingly the critical issue of non-stationarity appears to have been significantly overlooked in the literature on COVID-19. Nonetheless, as we show in the following, save few creditable exceptions, the reporting rate has generally increased substantially as the outbreak was gathering momentum, along with the stepping up of the number of tests conducted. Let us stress this point once more – any modelling efforts will be pointless unless non-stationarity of reporting is properly accounted for.

While infected and recovered bear the marks of the convolution with unpredictably non-stationary reading frames, it can be argued that the time series of deceased individuals provide a more reliable picture of the true, bias-free underlying dynamics of the disease. In fact, while there might have been inaccuracies in attributing deaths to COVID-19, it can be surmised that these should lead to systematic and reasonably stationary errors.

Contrary to intuition, and to an excellent degree of accuracy, a death-only analysis of the data shows that COVID-19 outbreaks in all countries fall in the simple SIR universality class, mean-field compartmental models originated by the the seminal 1927 paper by Kermack and McKendrick.^13^ Importantly, this fact can only be brought to the fore in the reduced space of deaths and their time derivatives, as this allows one to filter out virtually all spurious effects associated with non-stationary reading frames, which have been systematically underestimated in the vast majority of reports.

Taken together, our analysis suggests that simple mean-field models can be meaningfully invoked to gather a quantitative and robust picture of the COVID-19 epidemic spreading. Additionally, as a natural byproduct of our reasoning, we were able to calculate the time-dependent reporting fractions for infected and recovered individuals. These provide a clear a-posteriori reading frame for the relative position of the true epidemic peak and that observed from the reported data, which is found to be typically shifted away artificially in the future. Furthermore, casting the SIRD dynamics as a function of the deaths time series and its derivative leads us to derive simple formulae that prove invaluable for real-time monitoring of the disease. As an important corollary of our analysis, doubts can be cast on all procedures targeted at estimating the reproduction index of the epidemics based on the *raw* time series of infected (and recovered), i.e. not corrected for non-stationary testing.

## How to look at the reported figures

Within a simple mean-field compartmental model, people are divided into different *species*.^14^ For example, in the susceptible (S), infected (I), recovered (R), dead (D) scheme (SIRD),^15^ any individual in the fraction of the overall population that will eventually get sick belongs to one of the aforementioned classes. The latter represent the different stages of the disease and their size is used to model its time course. Let *S_0_ = S*(0) be the size of the initial pool of susceptible individuals. Then the SIRD dynamics is governed by the following set of coupled differential equations

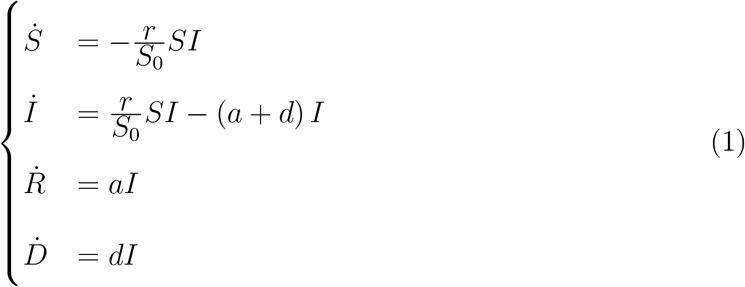

where *Ẋ* stand for the time derivative of *X = S, I, R, D*. The parameter *r* represents the infection rate, i.e. the probability per unit time that a susceptible individual contracts the disease when she enters in contact with an infected person. The parameters *a* and *d* denote the recovery and death rates, respectively. Note that, *S(t)* + *I(t)* + *R(t)* + *D(t) = S*_0_ + *I*_0_ is an invariant of the SIRD dynamics, i.e. it remains constant as time elapses^1^ In general, one could attempt to calibrate model (1) against the available time series for the infected, recovered and deaths, so as to determine the best-fit estimates of *r*, *a*, *d* and *S*_0_. However, it may be argued that this procedure amounts to over-fitting, due to the intrinsic high correlation between *r* and *S*_0_ in the model. More commonly, a face-value figure for *S*_0_ is assumed from the start, e.g. the size of a country or of some specific region. However, this choice is somehow arbitrary, as the *true* basin of the epidemics can in principle be determined meaningfully only a posteriori.

More importantly, as argued above, there is a more fundamental limitation to any attempt of adjusting a model to the raw figures beyond specific, model-dependent issues. The cause is the time-varying bias in the reported data. Essentially, this translates into an a-priori unknown discrepancy between the true size of infected and recovered and the figures released officially. The key observation at this point is that the time series of deaths, *D*(*t*), can be regarded as the only data set that returns a faithful representation of the epidemics course. More precisely, it appears reasonable to surmise that all unknown sources of errors on deaths can be considered as roughly stationary over the time span considered.

It is well-known that the SIR dynamics can be recast in a particularly appealing form in the space (*Ṙ*, *R*), *R* denoting in this case the overall class of recovered and deceased.^16,17^ When one distinguishes between the two populations *D* and *R* (the recovered), as in the SIRD dynamics (1), a similar manipulation leads to (see Methods)

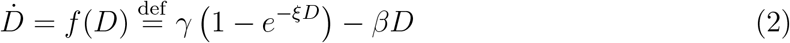

where *γ* = *dS*_0_, *ξ = r/(dS*_0_) and *β* = *a* + *d*. The differential equation (2) is equivalent to the full set of equations (1) and the associated conservation law. Overall, it provides an expedient and virtually bias-free means of testing whether the time evolution of an outbreak is governed by a SIRD-like dynamics.

Fig. 1 shows very clearly that the COVID-19 outbreak falls in the SIRD universality class. The data reported by national authorities in different countries are plotted in the plane (*Ḋ*, *D*) and compared with three-parameters fits performed with Eq. (2). The quality of the fits appears remarkably good, irrespective of the country and the actual severity of the epidemics in terms of deaths and deaths per day. Residual oscillations about the mean-field interpolating trends are visible, pointing to interesting next-to-leading order corrections, which would deserve further attention.

**Figure 1:**
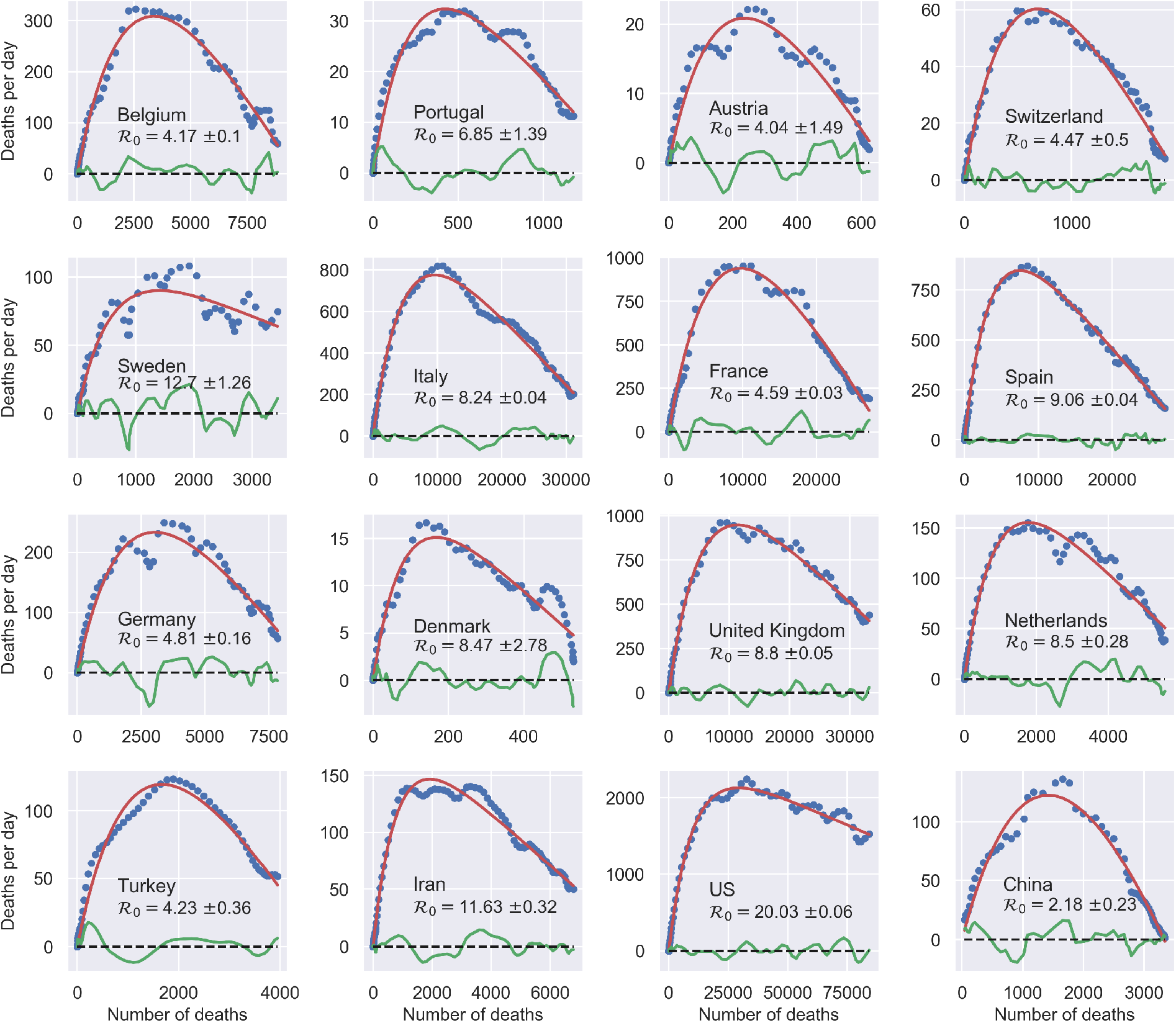
Universal behaviour of the COVID-19 outbreak in the (*Ḋ*, *D*) plane. When analysed in this space, outbreaks in different countries are described by the same three-parameters function *f(D*), Eq. (2). The reported values of the effective reproduction number at *t* = 0, 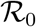 are computed as 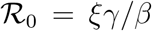. The data for *Ḋ* are centred rolling averages performed over a window of 6 days. Residuals most likely contain the (possibly time-shifted) blueprints of specific containment measures.^18^

It appears remarkable that a process that is inherently complex and unpredictably many-faceted can be accurately described in terms of an effective, highly simplified mean-field scheme, where many factors are deliberately omitted, such as space and different sorts of population stratification. Of course, the effectiveness of SIR schemes and simple modifications thereof are well-known.^19,20^ For example, a recent work shows that a modified SIR model where the infection rate is let vary with time can be employed to infer change points in the epidemic spread to gauge the effectiveness of specific confinement measures in Germany. ^18^ In an earlier work, two of the present authors had considered a similar approach to elaborate predictions of the outbreak in Italy that proved fundamentally right when compared with the evolution observed afterwards.^14^ What Fig. 1 critically adds to the picture is a bias-free confirmation that simple mean-field schemes can be used meaningfully to draw quantitative conclusions.

The best-fit values of the reproduction number 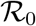 extracted from the above analysis should be considered as *effective* estimates of the overall severity of the outbreak in different countries. In particular, it can be argued that these should incorporate all sources of non-stationarity left besides the time-dependent bias associated with the evolution of the testing rate. Most likely, these amount to different degrees of (time-dependent) reduction of the infection rate *r*, following the containment measures. Interestingly, we find that, rather generally, the evolution of such non-stationary SIRD dynamics falls into the same universality class in the plane (*Ḋ*, *D*) as signified by *f*(*D*), Eq. (2) (see also section 3 in the supplementary material). More precisely, it turns out that a reduction of *r* by a factor *f* < 1 over a certain time window simply corresponds to a proportional rescaling of the reproduction number to a lower value, 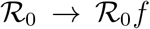. A similar conclusion holds if the non-stationarity causes the opposite effect, that is, *f* > 1. This might be the case, for example, of overlooked hotbeds or large undetected gatherings that cause the infection to accelerate at a certain point in time. Remarkably, whatever the direction, it can be proved that this rescaling is independent of the typical time scale over which the transition *r* → *fr* unfolds, as well as the point in time that marks its onset (see supplementary material).

An important corollary of our analysis is that the observed epidemic peak displayed by the time series of the *measured* infected, *I_M_(t*) typically occurs later than the true peak, the more so the more the testing rate is ramped up during the outbreak. The correct occurrence of the peak and a comprehensive understanding of how the testing activity controls the apparent evolution of the epidemics can be obtained with the following simple argument. In general, one can define two time-dependent *reporting fractions*, gauging the unknown disparity between the true and the measured populations

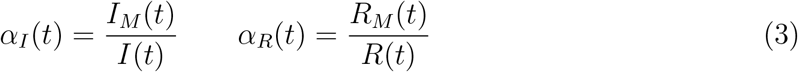

where *R_M_(t*) refers to the reported population of recovered persons. Taking into account the first definition in Eqs. (3), differentiation of the last equation in (1) with respect to time gives

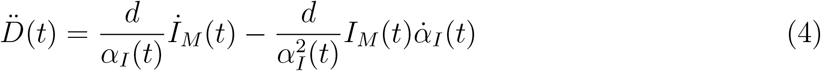

The apparent epidemic peak occurs when *İ_M_(t) =* 0, whereas we know from the fact that the epidemics is governed by a SIRD-like dynamics that the true peak occurs when 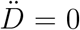, i.e. when the number of deaths/day reaches a maximum. Thus, from Eq. (4) one immediately sees that whether the apparent peak is observed before or after the *true* peak depends on the sign of the rate of reporting, 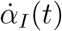. More precisely, if the testing activity is steadily ramping up (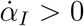), the true peak will occur earlier than the apparent one. This is because the reported infected will have a maximum for 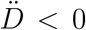, i.e. past the maximum of the *Ḋ*. Conversely, if the testing rate decreases, this will anticipate the apparent peak, giving the false impression that the worst might be over while the true number of infected is in fact still increasing. We find that this analysis applies to all countries considered in this paper (see also supplementary material), whereby either the former or the latter scenarios are invariably observed.

Fig. 2 illustrates the above considerations for the outbreak dynamics in three representative countries, each displaying one of the two characteristic behaviors. Technically, in order to compute the reporting fractions *α_I_*, *α_R_*, one needs to know the mortality *m* = *d*/*β* = *d*/(*a+d*). A good country-independent estimate for *m* has been reported recently by looking at the infinite-number of test extrapolation,^21^ which yields *m* = 0.012. Hence, combining Eqs. (1) with the definitions (3), it is not difficult to obtain

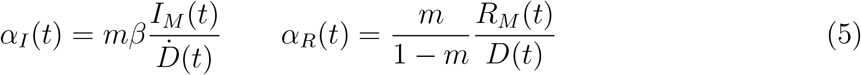

where *β* is estimated from the fits of the SIRD model in the plane *(Ḋ, D)*. Direct inspection of Fig. 2 confirms the above reasoning on the relative position in time of the apparent and true peaks. For example, it can be seen that tests have increased exponentially in Italy, starting from the end of March, which caused the apparent peak to occur nearly a month after the true one. Conversely, the testing activity has been vigorous in Germany at the onset of the outbreak, but appears to have slowed down substantially as the epidemics progressed. As a consequence, the apparent peak has been reported while the epidemics was still growing. In both cases, our estimate is consistent with the fact that only 15 % of infected persons are effectively reported. China seems to provide an instance of the marginal case 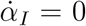. Despite an early ramping up, the reporting fraction of infected appears to be remarkably stable, which is consistent with the co-occurence of the true and the apparent peaks. According to the data, the testing activity has been resolute since the beginning (only about half of the infected eluding tracing), and has culminated with a scenario compatible with virtually all infected individuals being traced.

**Figure 2:**
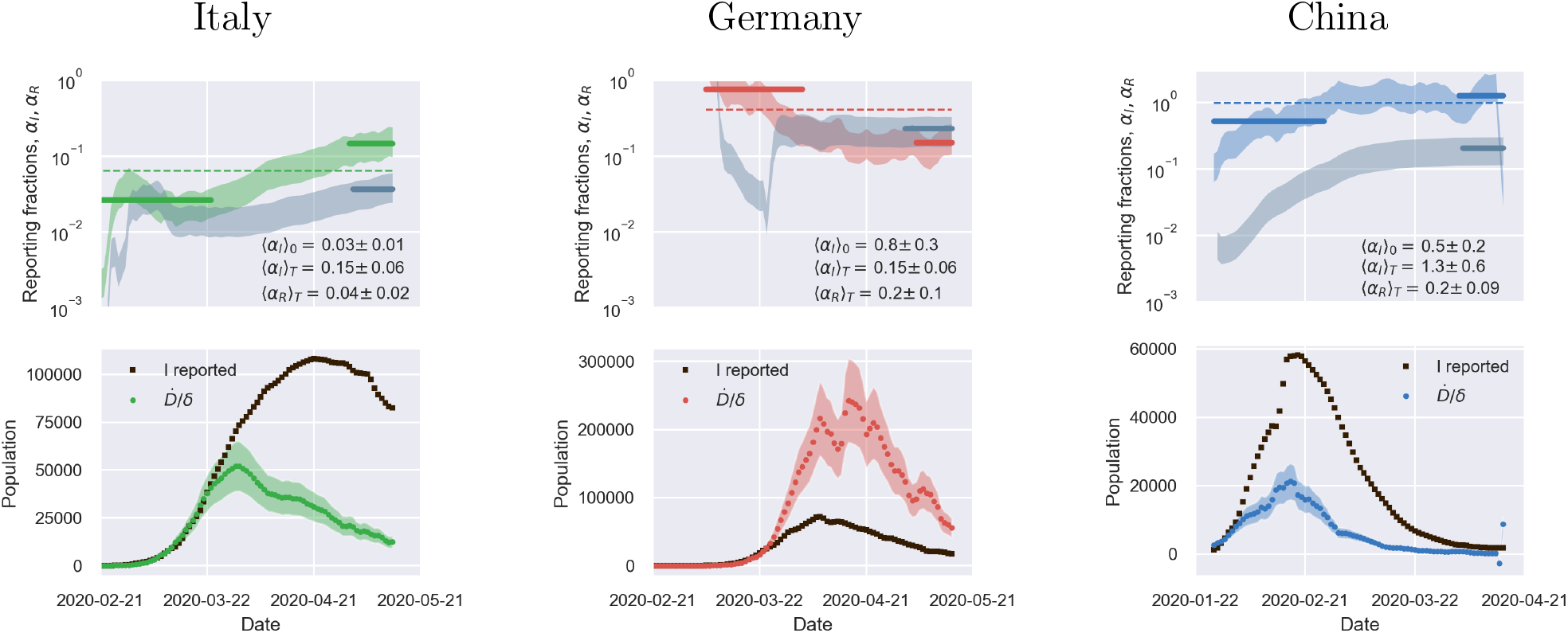
The derivative of the reporting fraction of infected controls the apparent epidemic peak. Top panels: fraction of true recovered reported, *α_R_(t)* (grey shaded regions) and fraction of true infected reported, *α_R_(t)*, computed as described in the text. The shaded regions correspond to an assumed mortality *m =* 0.012 ± 0.005.^21^ The thick lines mark averages on the last 15 % portions of the data (〈*α_I,R_*〉*_T_*), and first 40 % (〈*α_I_*〉_0_), while the dashed lines denote the overall average values of *α_I,R_* computed over the whole available time span. Bottom panels. The reported infected are compared to the rescaled deaths/day, 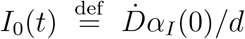, i.e. the infected that would have been reported if the reporting fraction had remained constant at its initial value *α_I_*(0). Technically, *δ = d/α_I_*(0) is obtained as the slope *δ* of a linear fit of *Ḋ* vs in the very early stage of the outbreak. The shaded regions correspond to the least-square errors found on 5. As soon as the testing rate starts varying appreciably, the two trends *I*_0_(*t*) and *I_M_*(*t*) diverge. If the testing rate is ramped up, the infected reported seem to flag an increase in the outbreak severity (Italy). Conversely, if the testing rate slows down (Germany), the reported infected underestimate the true peak. In particular, in this case the peak can be alarmingly anticipated. China shows a marginal behaviour, where the two peaks coincide, consistently with a remarkably stable reporting fraction of infected.

The calculations reported in Fig. 2 also confirm that counting instances of recovery is a much subtler task. Typically, countries seem to have needed some *adjustment* time to set up robust counting protocols, as it is apparent from the large fluctuations that are often observed in the trends of *α_R_* at early times (see also supplementary material). Another interesting feature that emerges clearly from the comparison of *α_I_* and *α_R_* is the average recovery time, which can be inferred for example by direct inspection of the testing activity in Italy. The curve *α_R_(t)* appears to be shifted in the future by about 10 days with respect to *α_I_*(*t*) which is consistent with the present consensus estimate of an average time for recovery of two weeks.

Our analysis shows how to pinpoint the true peak of the epidemics - the maximum of the deaths/day time series, *Ḋ*. It is thus natural to investigate how the different true peaks compare among different countries once these time series have been brought to a common time origin. This is illustrated in Fig. 3 for a number of European countries. It can be clearly appreciated that the apogee of the outbreak in different countries does not occur later than about one month after the first ten deaths reported. It is interesting to observe that in countries where the testing rate was vigorous at the onset of the outbreak (large (〈*α_I_*〉)_0_), e.g. Austria, Switzerland, Belgium, Germany^2^, the peak seems to occur earlier with respect to countries that have started ramping up their testing capacity later into the outbreak, such as Italy, Spain, France and U.K. (see supporting information). This might just have the obvious explanation that early tracking of infected individuals enables more effective isolation from the start, thus leading to a less severe evolution of the outbreak as there simply are less contagious people able to propagate the disease. It is also interesting to observe that peak position and height show some correlation with the initial reproduction numbers measured in the (*Ḋ*, *D*) plane (see again Fig. 1 and Tables S1 and S2 in the supplementary material). With some notable exceptions (again Netherlands and Denmark), countries that totalised higher deaths/day at peak tend to be characterised by higher values of 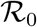. Overall, effective and timely testing seems to have been the key to successful containment strategies, as it is by now widely advocated,^22^ as opposed to muscular lockdowns imposed later into the outbreak.

**Figure 3:**
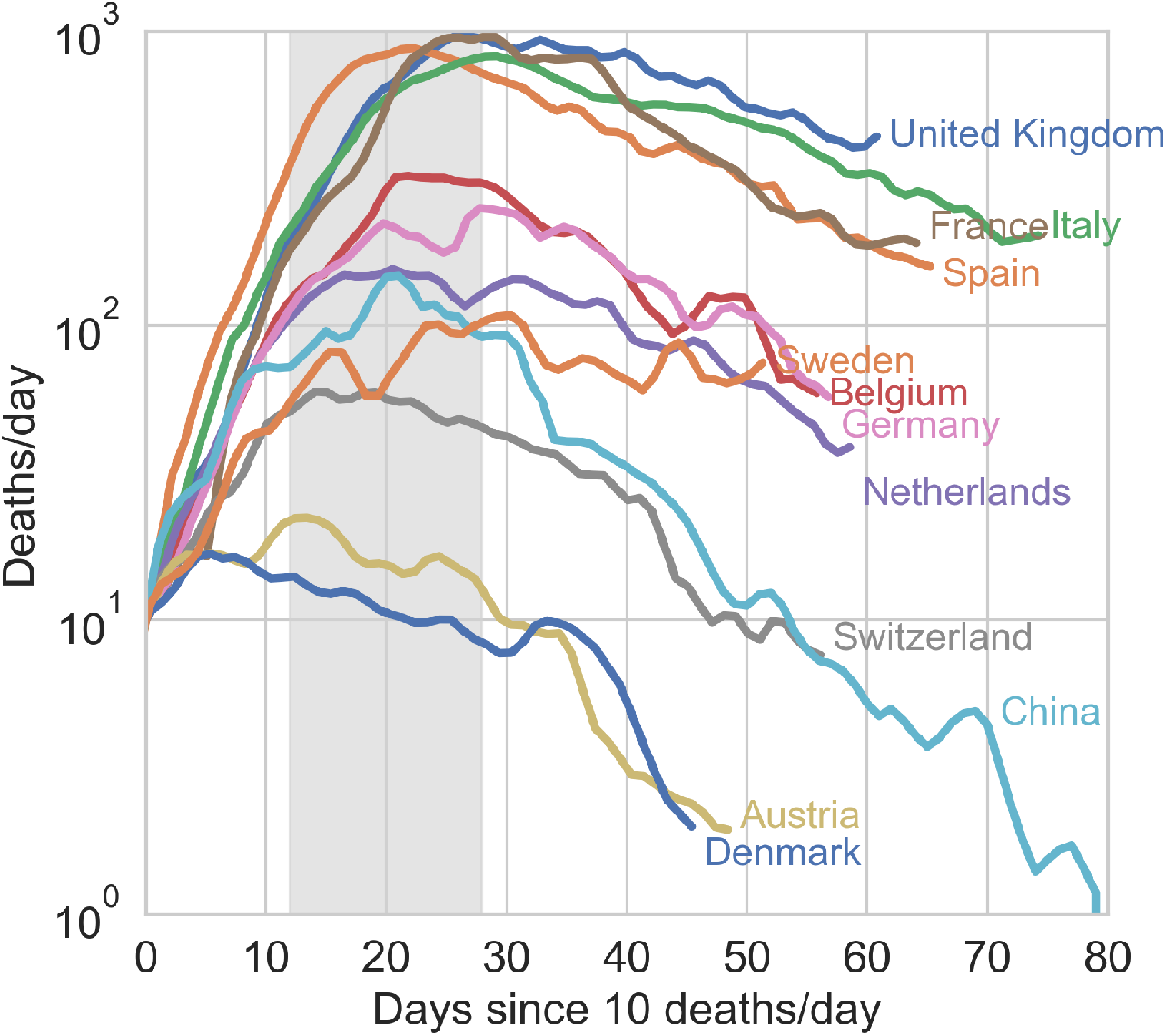
The true epidemic peak occurs within 30 days after the first ten deaths, the earlier the higher the initial testing rate. When shifted so that they coincide at the day where ten new deaths were first reported, the *Ḋ* time series reveal that the true epidemic peak occurs inevitably rather early in the outbreak, visibly not later than one month after the first ten casualties reported. Earlier vigorous testing is obviously key in containing the outbreak (see also discussion in the text).

## Summary and discussion

In summary, the observed behavior of the early 2020 COVID-19 outbreak has raised many diverse and puzzling issues in different countries. Many of them have adopted drastic containment measures, which had some effect on the spread of the virus but also left large sectors of the population world-wide grappling with the daunting perspective of pernicious economic recessions. On their side, scientists from all disciplines are having a hard time identifying recurring patterns that may help design and tune more effective response strategies for the next waves. In this letter we have shown that the spread of the epidemics caused by this elusive^4^ pathogen falls into the universality class of SIR models and extensions thereof. Based on this fact, an important corollary of our analysis is that early testing is not only key in making sense of the reported figures, but also in controlling the overall severity of the outbreak in terms of casualties.

It is intrinsically difficult to disentangle the effects imposed by containment measures from the natural habits of a given community in terms of social distancing. For example, Italy is recording three times as many deaths per inhabitant than Switzerland as we write. However, while it is true that Switzerland seemed to have tested nearly ten times as much initially (see supplementary material), it is also true that that country has never been put in full lockdown as it was decided in Italy. At the same time, Swiss people are known for their law-abiding attitude and social habits that make (imposed or suggested) social distancing certainly easier to maintain than in southern countries such as Italy.

The key message of this letter is that simple models should not be dismissed a priori in favour of more complex and supposedly more accurate schemes, which unquestionably go into more detail but at the same time rely necessarily on a large number of parameters that it is hard to fix unambiguously. All in all, we can state with certainty that vigorous early testing and accurate and transparent self-evaluation of the testing activity, assuredly virtuous attitudes in general, should be considered all the more a priority in the fight against COVID-19.

## Data

The data used in this paper span the interval 1/22/20 - 5/16/20 and have been retrieved from the github repository associated with the interactive dashboard hosted by the Center for Systems Science and Engineering (CSSE) at Johns Hopkins University, Baltimore, USA.^23^

## Methods

In this section we derive Eq. (2) and review the definition of the time-dependent reproduction rate 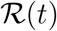. Combining the first and last equation in (1) we get

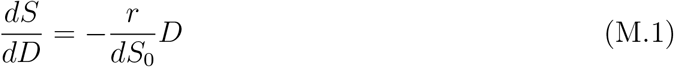

which can be readily integrated with the initial condition *D*(0) = 0, giving

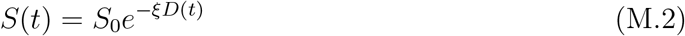

with *ξ* = *r/(dS*_0_). Furthermore, since *R*(0) = 0, one has from Eqs. (1) that *R(t) = (a/d)D(t)*. Taking into account that by definition *I(t) = Ḋ/d*, the conservation law, *S(t*) + *I(t)* + *R(t*) + *D(t) = S*_0_ + I_0_, can be recast in the following form

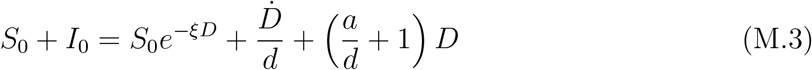

Multiplying through by *d* and taking into account that 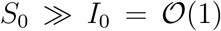, yields directly Eq. (2) with

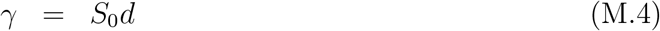

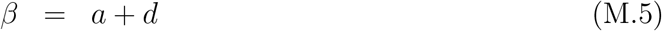

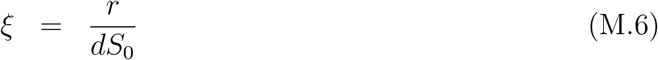

We now turn to discussing the definition of 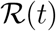. The second equation in (1) can be rewritten as

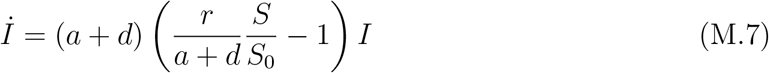

Following standard convention, we define the time-dependent reproduction number as

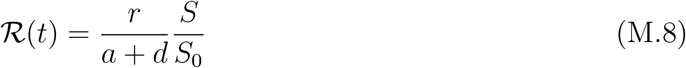

which, for *t* = 0, yields (see Eqs. (M.4), (M.5), (M.6))

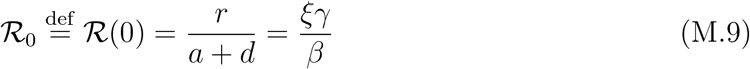

From (1) one readily obtains

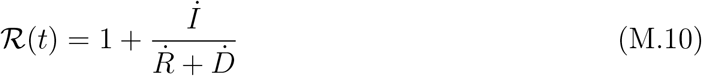

an expression which can be in principle employed to track the evolution in time of the epidemics. Recall, however, that only *I_M_(t*) and *R_M_(t*) are accessible to direct measurements. The associated reporting fractions are not known a priori and this may severely bias the analysis if the reported figures are used in Eq. (M.10) or extensions thereof. To overcome this limitation, it is expedient to combine the definition (M.8) with Eq. (M.2) to obtain a closed expression for 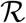 that only depends on *D*, namely

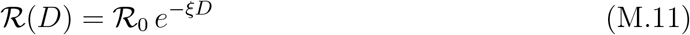

Eq. (M.11) can be used to estimate the time evolution of the reproduction number without the bias introduced by the non-stationarity of the reporting activity. In the supplementary material, it is shown that the curve of the infected *I* peaks at 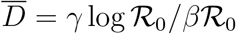. Plugging the latter expression into the right-hand side of Eq. (M.11) yields 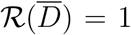, as it should for consistency requirements.

## Data Availability

The data have been retrieved by the public repository of Johns Hopkins University on GitHub

## Supplementary information

### Infection peak

In this section we derive some useful relations about key quantities at the infection peak, namely the time 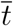 at which the number of infected persons, *I(t*), reaches its maximum value, 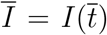. This can be determined from the condition 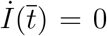. From the fourth equation of (1) in the main text, we get 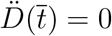, then using the definition of *f* (*D*) we can write

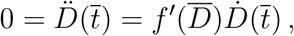

where 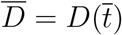. Being D a monotonic increasing function we have, *Ḋ(t*) > 0 for all *t*, hence the latter condition is equivalent to

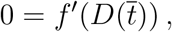

where the’ stand for the derivative of *f* with respect to the variable *D*. This equation can be straightforwardly solved to give

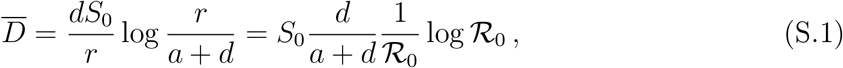

where 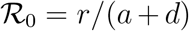 (see also Methods in the main text). We are now able to compute the remaining key quantities at the peak time 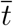. Indeed, the relations developed in the Methods section in the main text imply

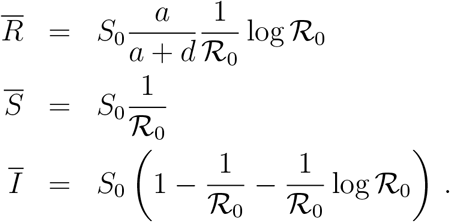

Let us observe that these quantities, as well as 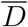, scale proportionally to the system size, *S*_0_, as one can appreciate from Fig. 4 where we report 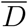 as a function of *S*_0_. The latter is computed as *S*_0_ = *γ*/*d* (see main text). Here *d* = *δα_I_*(0), where *δ* is the slope of a linear fit *Ḋ* vs *I_M_* at short times and *γ* is a parameter of the function *f* (*D*) obtained from a fit in the plane *(Ḋ, D)* (see Fig. 1 in the main text).

**Figure 4:**
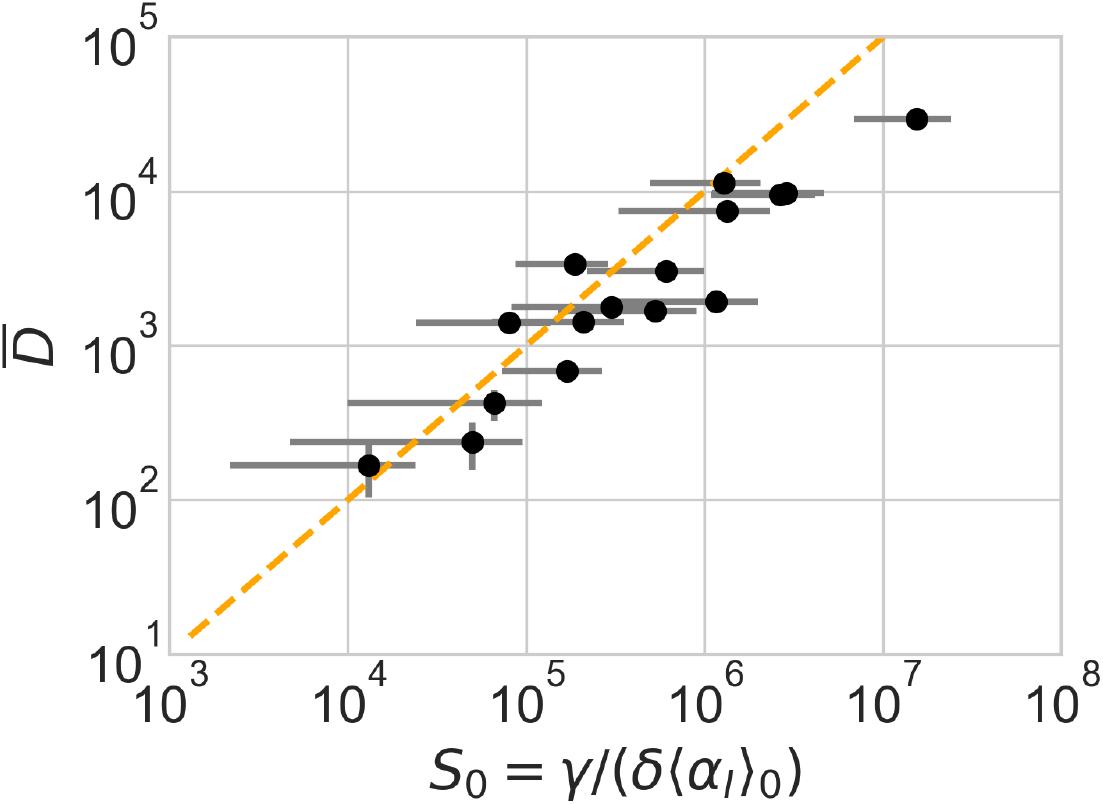
Number of deaths at the true infection peak vs effective population size. estimated self-consistently (see Table 2). A direct proportionality is observed, in agreement with relation (S.1). The vertical and horizontal bars denote the errors reported in Table 2, calculated according to standard error propagation from the formulae used to compute the two quantities. The orange dashed line is the arbitrary proportionality relation 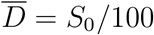, drawn to provide an order of magnitude of the ratio between deaths at peak and effective size of the infection basin.

**Table 1:** Best-fit values of the fitting parameters in the (*Ḋ, D)* plane *γ, β, ξ* and of the linear fit *Ḋ* = *δI_M_*. The errors reported have been computed by constructing confidence ellipses at 90 % confidence level (*γ, β, ξ*), and as standard least-squares errors for the linear fit to extract the slope *δ*.

| Country | $\gamma \pm \Delta\gamma$ (days<sup>-1</sup>) | $\beta \pm \Delta\beta$ (days<sup>-1</sup>) | $\xi \pm \Delta\xi$ | $\delta \pm \Delta\delta$ (days<sup>-1</sup>). |
| --- | --- | --- | --- | --- |
| Belgium | $737 \pm 6$ | $0.0751 \pm 0.0006$ | $0.000426 \pm 4 \times 10^{-6}$ | $0.013 \pm 0.002$ |
| Portugal | $56 \pm 3$ | $0.038 \pm 0.003$ | $0.0046 \pm 0.0003$ | $0.004 \pm 0.001$ |
| Austria | $51 \pm 6$ | $0.075 \pm 0.009$ | $0.0059 \pm 0.0008$ | $0.0012 \pm 0.0003$ |
| Switzerland | $136 \pm 5$ | $0.067 \pm 0.002$ | $0.0022 \pm 9 \times 10^{-5}$ | $0.0026 \pm 0.0005$ |
| Sweden | $125 \pm 2$ | $0.0177 \pm 0.0008$ | $0.0018 \pm 5 \times 10^{-5}$ | $0.009 \pm 0.002$ |
| Italy | $1245 \pm 2$ | $0.0333 \pm 0.0001$ | $0.0002206 \pm 5 \times 10^{-7}$ | $0.016 \pm 0.004$ |
| France | $2086 \pm 5$ | $0.0713 \pm 0.0001$ | $0.0001572 \pm 4 \times 10^{-7}$ | $0.006 \pm 0.001$ |
| Spain | $1312 \pm 2$ | $0.0426 \pm 0.0001$ | $0.0002946 \pm 6 \times 10^{-7}$ | $0.016 \pm 0.004$ |
| Germany | $500 \pm 5$ | $0.0537 \pm 0.0006$ | $0.000518 \pm 6 \times 10^{-6}$ | $0.001 \pm 0.0002$ |
| Denmark | $24 \pm 2$ | $0.036 \pm 0.004$ | $0.013 \pm 0.002$ | $0.005 \pm 0.001$ |
| U. K. | $1483 \pm 2$ | $0.0323 \pm 0.0001$ | $0.0001919 \pm 4 \times 10^{-7}$ | $0.019 \pm 0.005$ |
| Netherlands | $246 \pm 2$ | $0.035 \pm 0.0004$ | $0.00121 \pm 2 \times 10^{-5}$ | $0.012 \pm 0.003$ |
| Turkey | $283 \pm 8$ | $0.058 \pm 0.002$ | $0.00086 \pm 2 \times 10^{-5}$ | $0.0041 \pm 0.0009$ |
| Iran | $209 \pm 1$ | $0.0229 \pm 0.0002$ | $0.00128 \pm 1 \times 10^{-5}$ | $0.011 \pm 0.003$ |
| US | $2662 \pm 2$ | $0.0135 \pm 0.0001$ | $0.0001021 \pm 1 \times 10^{-7}$ | $0.006 \pm 0.001$ |
| China | $670 \pm 30$ | $0.168 \pm 0.005$ | $0.00055 \pm 2 \times 10^{-5}$ | $0.006 \pm 0.002$ |

**Table 2:** Number of deaths at the true infection peak, 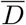, computed self-consistently as 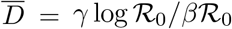 (see Eq. (S.1)) and estimated values for the *effective* basin of susceptible *S*_0_, computed as *S*_0_ = *γ/d = γ/(δ*〈*α_I_*〉_0_). The averages in 〈*a_I_*〉_0_ correspond to the first 40 % of the data (starting from 22/01/2020), see also Fig. 2 in the main text. A plot of 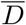 vs *S*_0_ is reported in Fig. 4.

| Country | $\bar{D} \pm \Delta\bar{D}$ | $S_0 \pm \Delta S_0$ |
| --- | --- | --- |
| Belgium | $3360 \pm 80$ | $200000 \pm 100000$ |
| Portugal | $400 \pm 100$ | $70000 \pm 60000$ |
| Austria | $240 \pm 80$ | $50000 \pm 50000$ |
| Switzerland | $680 \pm 70$ | $200000 \pm 100000$ |
| Sweden | $1400 \pm 200$ | $80000 \pm 60000$ |
| Italy | $9560 \pm 60$ | $3000000 \pm 2000000$ |
| France | $9710 \pm 70$ | $3000000 \pm 2000000$ |
| Spain | $7480 \pm 40$ | $1000000 \pm 1000000$ |
| Germany | $3000 \pm 100$ | $600000 \pm 400000$ |
| Denmark | $170 \pm 60$ | $10000 \pm 10000$ |
| U. K. | $11340 \pm 90$ | $1300000 \pm 800000$ |
| Netherlands | $1770 \pm 70$ | $300000 \pm 200000$ |
| Turkey | $1700 \pm 100$ | $500000 \pm 400000$ |
| Iran | $1920 \pm 70$ | $1200000 \pm 800000$ |
| US | $29300 \pm 100$ | $15000000 \pm 9000000$ |
| China | $1400 \pm 100$ | $200000 \pm 100000$ |

### Rescaled variables

In the main text we have shown (see Fig.1) that the epidemic spreading of the COVID-19 falls for a large number of countries in the SIR universality class, once read in the plane *(Ḋ*, *D*). Indeed, all the data fit very well on the curve *Ḋ* = *f* (*D*), the latter depending on three parameters. One can go one step forward and show that there is a particular choice of rescaled variables such that all the curves *Ḋ* = *f* (*D*) collapse on a one-parameter family of curves, indexed by the effective basic reproduction number, 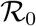. Let the define a new variable x and a new time *t′* by

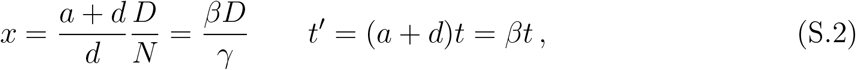

where *N* = *S*_0_ + *I*0 ≈ *S*_0_. Then (see Eq. (2) in the main text and Methods)

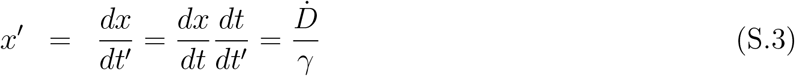

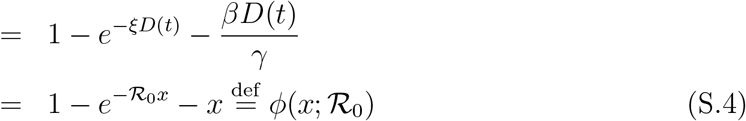

where we have used the relation 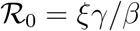. The universal function is unimodal, 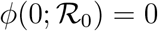 for all values of 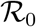 and there is a single positive root, 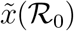, which converges to 1 for 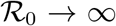. The function *ϕ* is linear for small values of *x* with slope 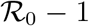, while at 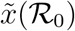 one has

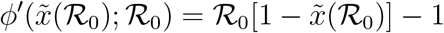

It is easy to see that 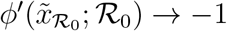 as 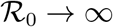.

Fig. 5 reports the same data as plotted in Fig. 1 in the main text, but in the rescaled plane (*x*′, *x*). It is apparent that there exist two main clusters of outbreaks, one with values of 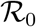 between 4 and 5 and one with values of 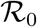 between 8.5 and 9.5. The outbreak in China is the only observed case where 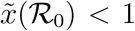, due to the low value of 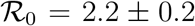 computed from the fit.

**Figure 5:**
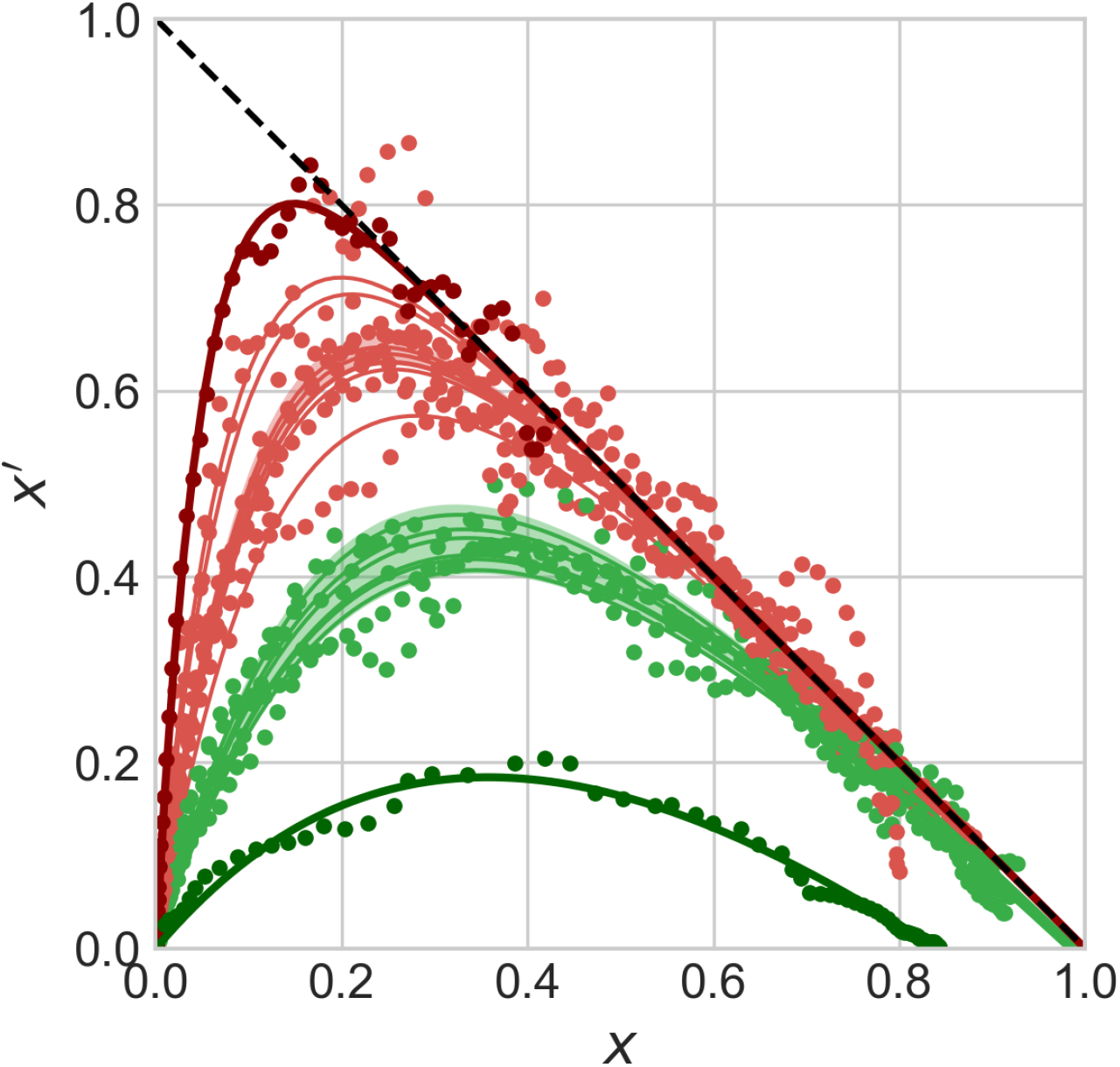
Universal behaviour of the COVID-19 outbreak in the rescaled plane (*x′,x*). Symbols stand for the data *Ḋ*, *D* rescaled according to the prescriptions (S.3) and (S.2), respectively. The green and red shaded regions highlight the intervals 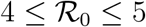 and 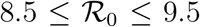, respectively. The thick green and red lines denote the data from the outbreaks in China (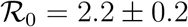) and the United States (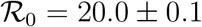), respectively.

### Non-stationary reduction of the infection rate and the effective reproduction number

The reproduction numbers measured from the fits performed in the (*Ḋ, D)* plane should be considered as *effective* values. These most likely incorporate all sources of non-stationarity left besides the time-dependent bias associated with the evolution of the testing rate. In particular, when imposed on the population, the effect of a lockdown is that of reducing over a characteristic (possibly rather short) period of time the rate of infection *r*. The question then arises whether the portrait in the (*Ḋ*, *D*) plane of a SIRD evolution where the rate of infection r is a time-dependent function will still be described by the same universal function *f* (*D*) but with an *effective* reproduction number 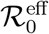. It turns out that the answer is in the affirmative and the rationale for this is deceptively simple.

In order to explore this question, we consider a modified version of the SIRD model (Eqs. (1) in the main text), where the infection rate *r* is let vary with time.^14^ More precisely, if some containment measures were enforced at time *t\**, causing their effect on a typical time ∆*t*, we may take

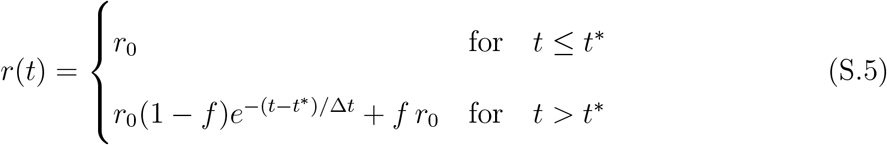

where *r*_0_ is the initial, pre-lockdown infection rate that defines the *true* initial reproduction number, 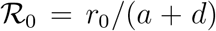. The fraction *f* gauges the asymptotic reduction (if *f* < 1) of the infection rate afforded by the containment measures. Of course, this kind of description can be easily adapted to the case where the non-stationarity causes the opposite effect, that is, *f* > 1. This might be the case, for example, of overlooked hotbeds or large undetected gatherings that cause the infection to accelerate on average at a certain point in time.

Whatever the effect of the non-stationarity introduced by *r(t)*, the effect in the rescaled plane *(x′, x)* turns out to be extremely simple and apparently robust with respect to the choice of the characteristic time scales *t*^*^, ∆*t* and also of the specific functional form that describes the reduction from *r*_0_ to *fr*_0_. This is illustrated in Fig. 6 for a choice of *f* in the interval [0.1, 3]. The evolution of outbreaks generated from a given *root* one, that is, one with a given infection rate *r*(0) = *r*_0_, can always be described by the same one-parameter function 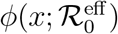 with an effective reproduction number 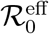 that only depends on *f*. Moreover 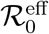 for *f* < 1 is simply the *t* = 0 value reduced by the same factor, that is

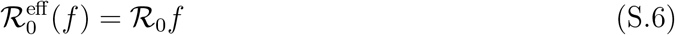

**Figure 6:**
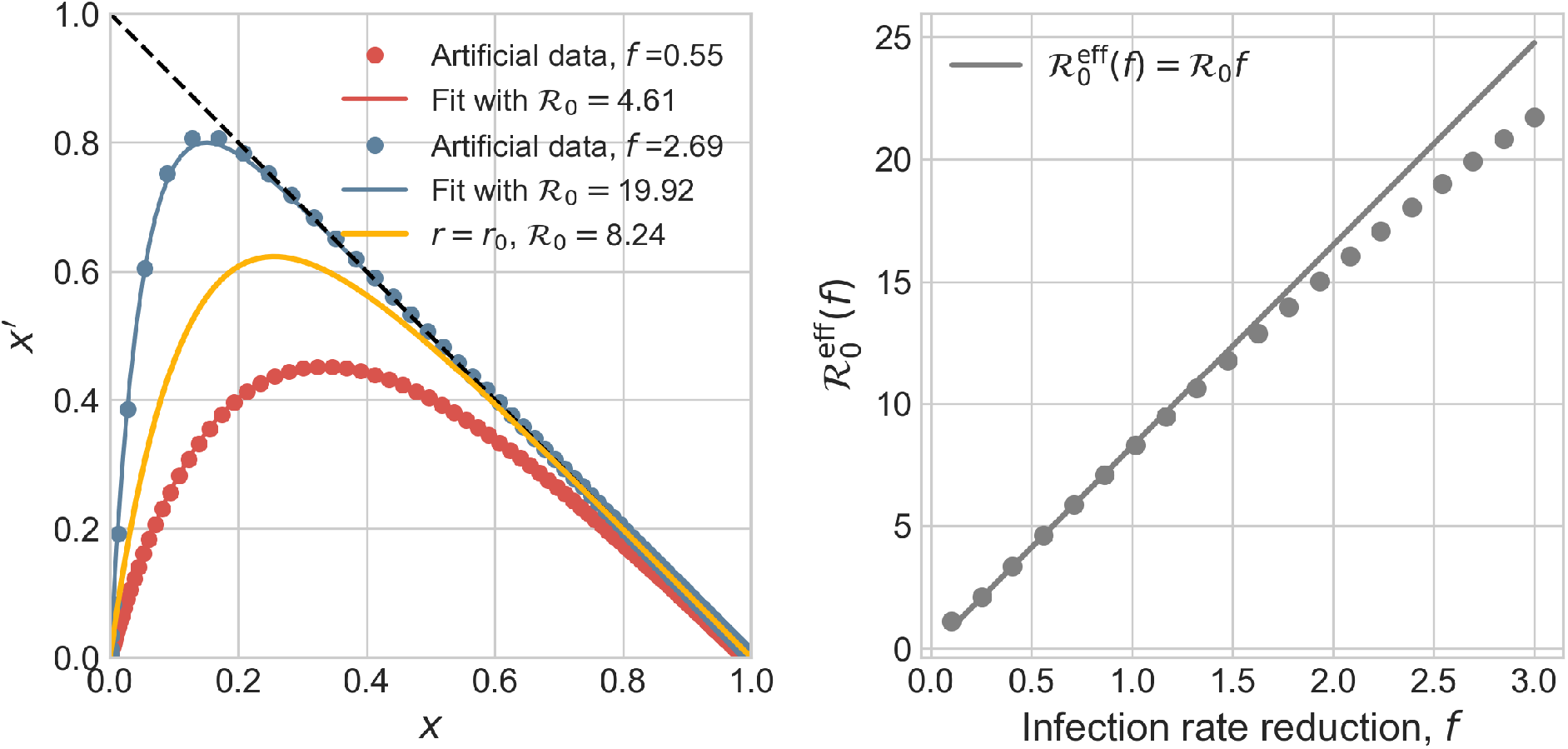
SIRD-like behaviour in the rescaled plane (*x′*, *x*) is observed for non-stationary infection rate with an effective reproduction number. Starting from a given set of parameters, *r*_0_ = 0.275 days^−1^, *a* = 0.0309 days^−1^, *d* = 0.00235 days^−1^, corresponding to 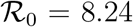, we have generated different *artificial* SIRD dynamics starting from the same initial conditions, *S*_0_ = 528747, *I*_0_ = 1 according to Eq. (S.5) with *f* ∈ [0.1, 3], *t*\* = 20 days and ∆*t* = 3.5 days. Each non-stationary SIRD dyamics has been fitted in the *(x′*, *x*) plane with the function 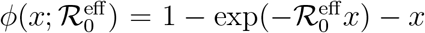 (two instances are plotted in the left panel). The effective reproduction numbers are plotted in the right panel against the corresponding reduction of infection rate, *f*. These seem to be described by the same universal linear reduction, 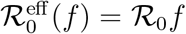, irrespective of the characteristic time scales that govern the non-stationarity and even of the specific function the drives the reduction from *r*_0_ to *fr*_0_ < *r*_0_ (see text).

Remarkably, this conclusion, for *f* < 1, does not depend on the particular choice of the time scale ∆*t*, nor on the onset time *t*^*^, nor on the particular choice for the function that describes the fading of the infection rate. For example, a function of the kind 1/(1 + *x*) instead of an exponential damping in Eq. (S.5) yields exactly the same result (S.6), that we might then consider a general fact.

**Figure 7:**
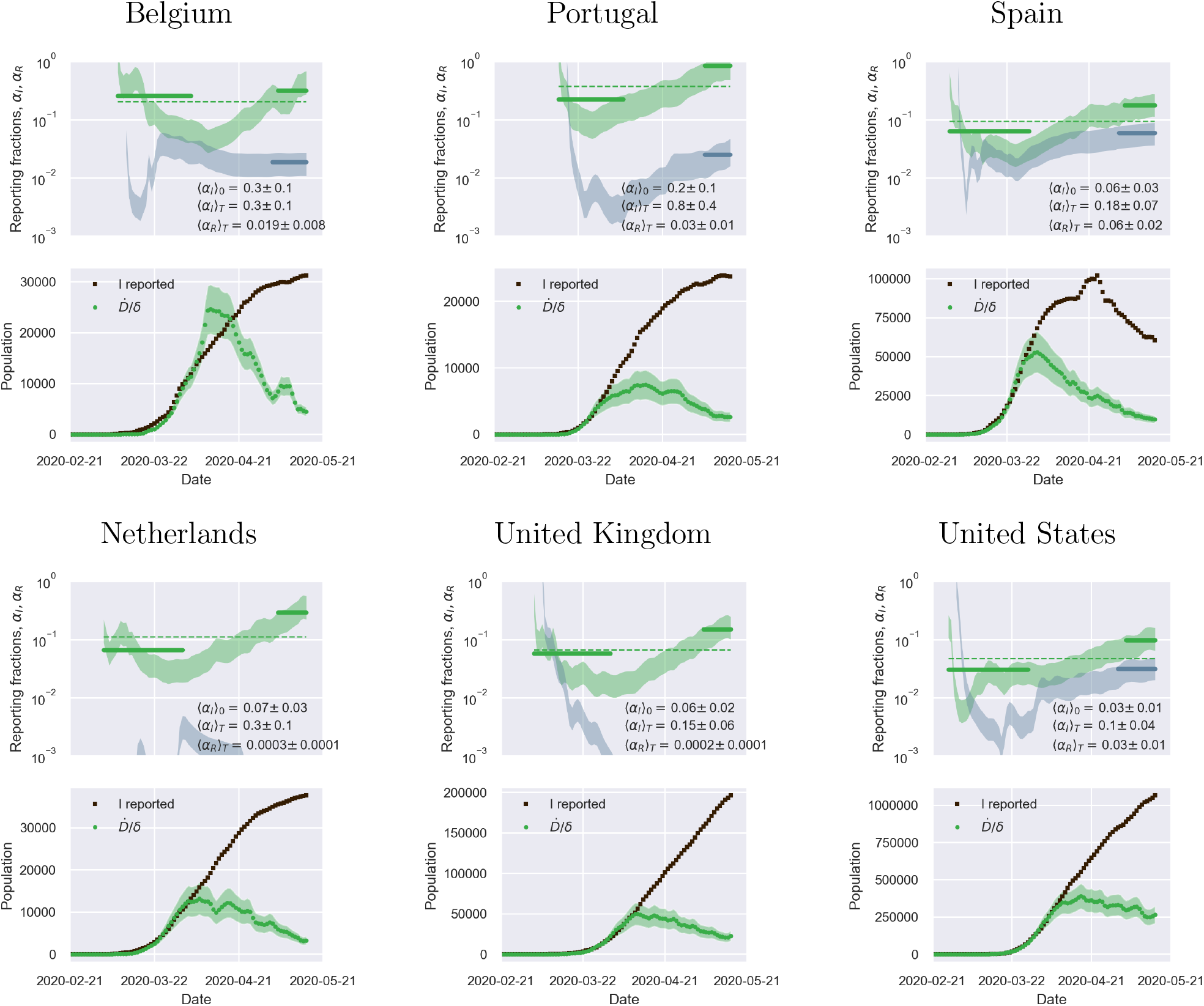
A positive derivative of the reporting fraction of infected causes a delay of the apparent epidemic peak with respect to the true one - I. Top panels: fraction of true recovered reported, *α_R_(t)* (grey shaded regions) and fraction of true infected reported, *α_I_(t)*, computed as described in the text. The shaded regions correspond to an assumed mortality *m =* 0.012 ± 0.005.^21^ The thick lines mark averages on the last 15 % portions of the data *((α_I,R_*)*_T_*), and first 40 % (〈*α_I_*〉_0_), while the dashed lines denote the overall average values of *α_I_,_R_* computed over the whole available time span. Bottom panels. The reported infected are compared to the rescaled deaths/day, 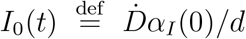, i.e. the infected that would have been reported if the reporting fraction had remained constant at its initial value *α_I_*(0). Technically, *δ = d/α_I_*(0) is obtained as the slope *δ* of a linear fit of *Ḋ* vs *I_M_* the very early stage of the outbreak. The shaded regions corresponds to the least-square errors found on *δ*.

**Figure 8:**
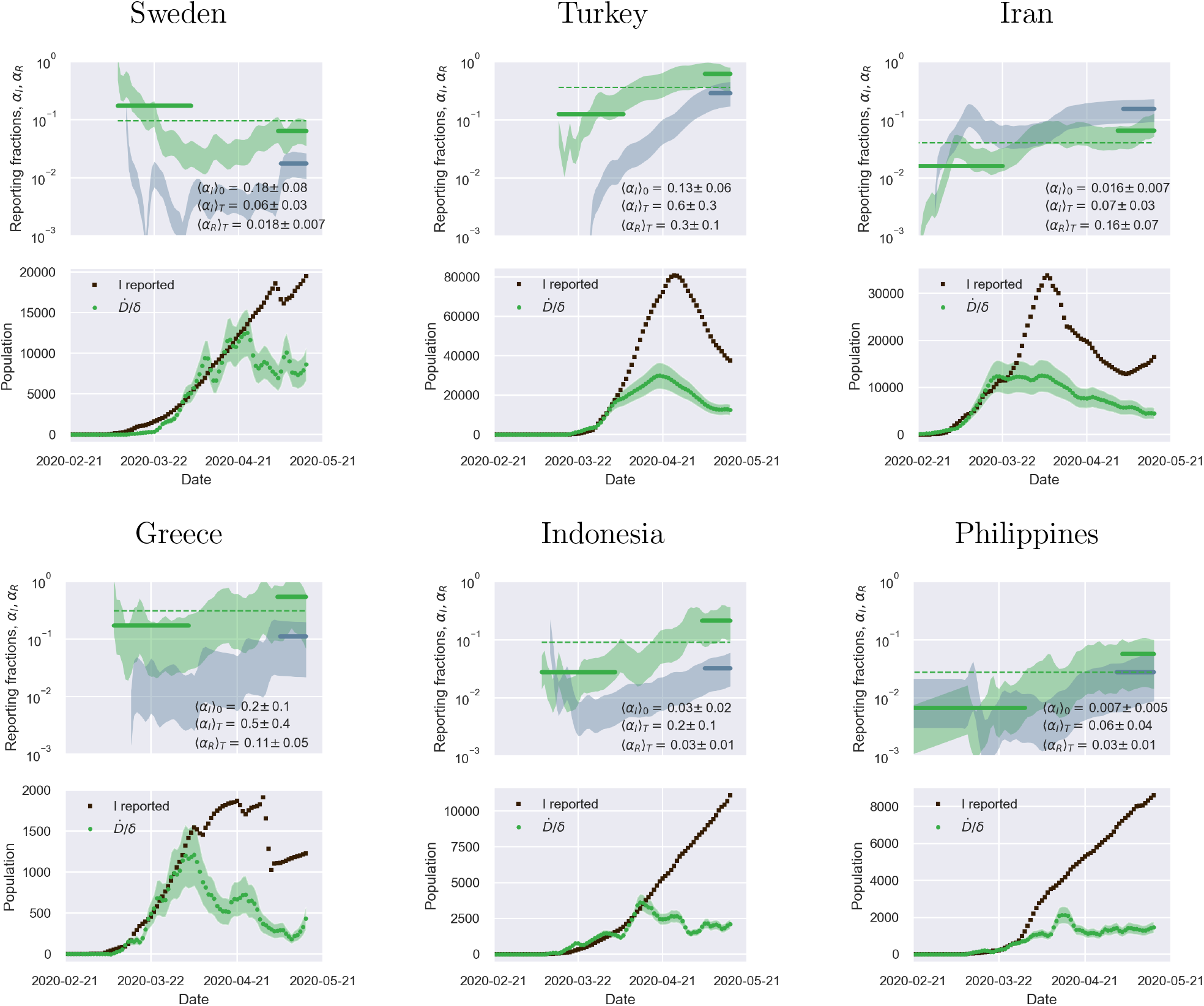
A positive derivative of the reporting fraction of infected causes a delay of the apparent epidemic peak with respect to the true one - II. Top panels: fraction of true recovered reported, *α_R_*(*t*) (grey shaded regions) and fraction of true infected reported, *α_I_*(*t*), computed as described in the text. The shaded regions correspond to an assumed mortality *m* = 0.012 ± 0.005.^21^ The thick lines mark averages on the last 15 % portions of the data (〈*α_I,_*_R_〉*_T_*), and first 40 % (〈*α_I_*〉_0_), while the dashed lines denote the overall average values of *α_I,R_* computed over the whole available time span. Bottom panels. The reported infected are compared to the rescaled deaths/day, 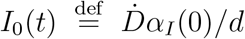, i.e. the infected that would have been reported if the reporting fraction had remained constant at its initial value *α_I_*(0). Technically, *δ* = *d/α_I_*(0) is obtained as the slope S of a linear fit of *Ḋ* vs *I_M_* in the very early stage of the outbreak. The shaded regions corresponds to the least-square errors found on *δ*.

**Figure 9:**
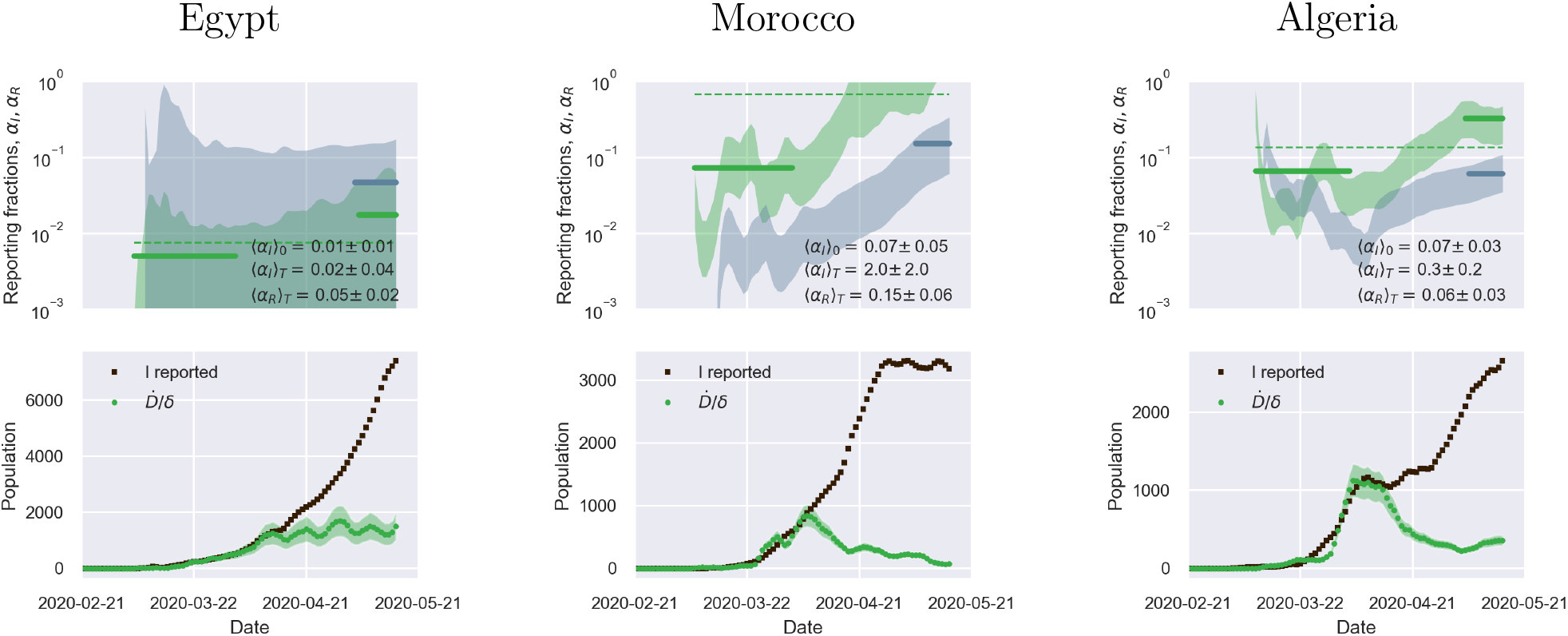
A positive derivative of the reporting fraction of infected causes a delay of the apparent epidemic peak with respect to the true one - III. Top panels: fraction of true recovered reported, *α_R_*(*t*) (grey shaded regions) and fraction of true infected reported, *α_I_*(*t*), computed as described in the text. The shaded regions correspond to an assumed mortality *m* = 0.012 ± 0.005.^21^ The thick lines mark averages on the last 15 % portions of the data (〈*α_I,R_*〉)*_T_*), and first 40 % (〈*α_I_*〉_0_), while the dashed lines denote the overall average values of *α_I_,_R_* computed over the whole available time span. Bottom panels. The reported infected are compared to the rescaled deaths/day, 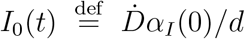, i.e. the infected that would have been reported if the reporting fraction had remained constant at its initial value *α_I_*(0). Technically, *δ* = *d/α_I_*(0) is obtained as the slope *δ* of a linear fit of *Ḋ* vs *I_M_* in the very early stage of the outbreak. The shaded regions corresponds to the least-square errors found on *δ*.

**Figure 10:**
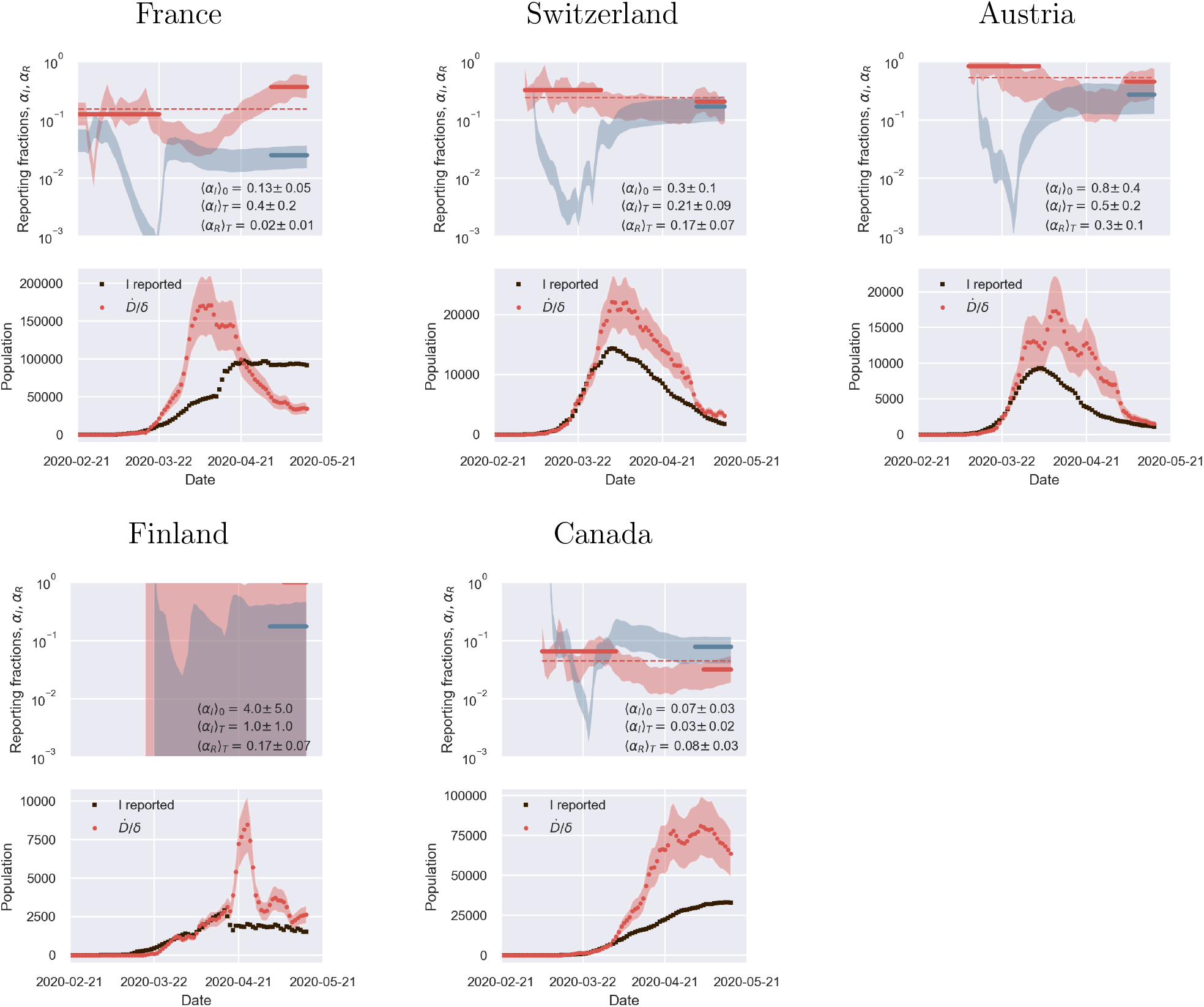
A negative derivative of the reporting fraction of infected deceptively anticipates the apparent epidemic peak with respect to the true one. Top panels: fraction of true recovered reported, *α_R_(t)* (grey shaded regions) and fraction of true infected reported, *α_I_(t)*, computed as described in the text. The shaded regions correspond to an assumed mortality *m* = 0.012 ± 0.005.^21^ The thick lines mark averages on the last 15 % portions of the data ((*α_I_*,*_R_*)*_T_*), and first 40 % *((α_I_*)_0_), while the dashed lines denote the overall average values of *α_I_,_R_* computed over the whole available time span. Bottom panels. The reported infected are compared to the rescaled deaths/day, 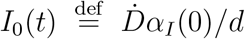, i.e. the infected that would have been reported if the reporting fraction had remained constant at its initial value *α_I_*(0). Technically, *δ* = *d/α_I_*(0) is obtained as the slope *δ* of a linear fit of *Ḋ* vs *I_M_* in the very early stage of the outbreak. The shaded regions corresponds to the least-square errors found on *δ*.

**Figure 11:**
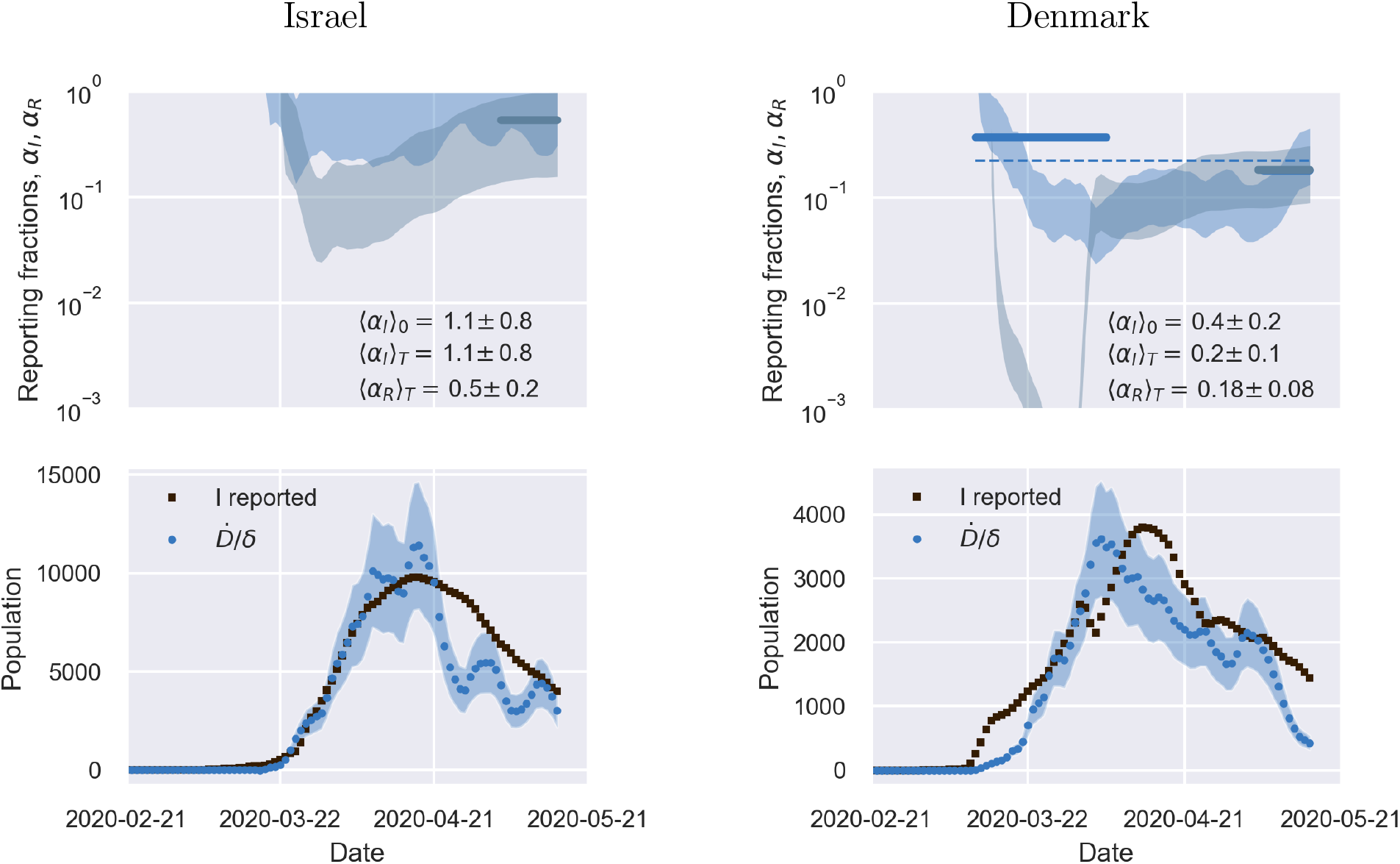
The outbreak in a reduced number of *marginal* cases is consistent with an overall flat reporting fraction of infected. Top panels: fraction of true recovered reported, *α_R_*(*t*) (grey shaded regions) and fraction of true infected reported, *α_I_*(*t*), computed as described in the text. The shaded regions correspond to an assumed mortality *m* = 0.012 ± 0.005.^21^ The thick lines mark averages on the last 15 % portions of the data *(〈*α_I_,_r_〉_t_), and first 40 % *(〈α_I_*〉_0_), while the dashed lines denote the overall average values of *α_I_,_R_* computed over the whole available time span. Bottom panels. The reported infected are compared to the rescaled deaths/day, 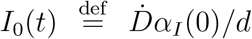, i.e. the infected that would have been reported if the reporting fraction had remained constant at its initial value *α_I_*(0). Technically, *δ* = *d*/*α_I_*(0) is obtained as the slope *δ* of a linear fit of *Ḋ* vs *I_M_* in the very early stage of the outbreak. The shaded regions corresponds to the least-square errors found on *δ*.

## Footnotes

1 It is reasonable to assume that *R*0) =0 as this coronavirus has never been in contact with humans before. Furthermore, 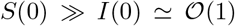, which justifies the way Eqs. (1) are written down (see also Methods).

2 Netherlands seem to be a remarkable exception to this observation (see supplementary material).

## References

(1) 2020; https://www.who.int/emergencies/diseases/novel-coronavirus-2019.

(2) Layne, S. P.; Hyman, J. M.; Morens, D. M.; Taubenberger, J. K. New coronavirus outbreak: Framing questions for pandemic prevention. Science Translational Medicine 2020, 12, eabb1469.

(3) Enserink, M.; Kupferschmidt, K. With COVID-19, modeling takes on life and death importance. Science 2020, 367, 1414–1415.

(4) Wang, Y.; Wang, Y.; Chen, Y.; Qin, Q. Unique epidemiological and clinical features of the emerging 2019 novel coronavirus pneumonia (COVID-19) implicate special control measures. Journal of Medical Virology 2020, 92, 568–576.

(5) Kupferschmidt, K. Why do some COVID-19 patients infect many others, whereas most don’t spread the virus at all? Science 2020,

(6) Gatto, M.; Bertuzzo, E.; Mari, L.; Miccoli, S.; Carraro, L.; Casagrandi, R.; Rinaldo, A. Spread and dynamics of the COVID-19 epidemic in Italy: Effects of emergency containment measures. Proceedings of the National Academy of Sciences 2020, 117, 1048410491.

(7) Giordano, G.; Blanchini, F.; Bruno, R.; Colaneri, P.; Di Filippo, A.; Di Matteo, A.; Colaneri, M. Modelling the COVID-19 epidemic and implementation of population-wide interventions in Italy. Nature Medicine 2020,

(8) Chinazzi, M. et al. The effect of travel restrictions on the spread of the 2019 novel coronavirus (COVID-19) outbreak. Science (New York, N.Y.) 2020, 368, 395–400.

(9) Balcan, D.; Colizza, V.; Gongalves, B.; Hu, H.; Ramasco, J. J.; Vespignani, A. Multiscale mobility networks and the spatial spreading of infectious diseases. Proceedings of the National Academy of Sciences 2009, 106, 21484–21489.

(10) Roda, W. C.; Varughese, M. B.; Han, D.; Li, M. Y. Why is it difficult to accurately predict the COVID-19 epidemic? Infectious Disease Modelling 2020, 5, 271–281.

(11) De Brouwer, E.; Raimondi, D.; Moreau, Y. Modeling the COVID-19 outbreaks and the effectiveness of the containment measures adopted across countries. medRxiv 2020, 2020.04.02.20046375.

(12) Hitchcock, C. In The Stanford Encyclopedia of Philosophy, summer 2020 ed.; Zalta, E. N., Ed.; Metaphysics Research Lab, Stanford University, 2020.

(13) Kermack, W. O.; McKendrick, A. G.; Walker, G. T. A contribution to the mathematical theory of epidemics. Proceedings of the Royal Society of London. Series A, Containing Papers of a Mathematical and Physical Character 1927, 115, 700–721.

(14) Fanelli, D.; Piazza, F. Analysis and forecast of COVID-19 spreading in China, Italy and France. Chaos, Solitons & Fractals 2020, 134, 109761.

(15) Anastassopoulou, C.; Russo, L.; Tsakris, A.; Siettos, C. Data-based analysis, modelling and forecasting of the COVID-19 outbreak. PLoS ONE 2020, 15, e0230405.

(16) Murray, J. D. Mathematical Biology; Springer, 1989.

(17) Vattay, G. Forecasting the outcome and estimating the epidemic model parameters from the fatality time series in COVID-19 outbreaks. 2020.

(18) Dehning, J.; Zierenberg, J.; Spitzner, F. P.; Wibral, M.; Neto, J. P.; Wilczek, M.; Priesemann, V. Inferring change points in the spread of COVID-19 reveals the effectiveness of interventions. Science 2020,

(19) Brauer, F.; Castillo-Chavez, C. Mathematical Models in Population Biology and Epidemiology, 2nd ed.; Springer, 2012.

(20) Diekmann, O.; Heesterbeek, J. A. P. Mathematical Epidemiology of Infectious Diseases: Model Building, Analysis and Interpretation; Wiley, 2000.

(21) Bastolla, U. How lethal is the novel coronavirus, and how many undetected cases there are? The importance of being tested. medRxiv 2020,

(22) Peto, J. Covid-19 mass testing facilities could end the epidemic rapidly. British Medical Journal 2020, 368, m1163.

(23) Dong, E.; Du, H.; Gardner, L. An interactive web-based dashboard to track COVID-19 in real time. The Lancet Infectious Diseases 2020, 3099, 19–20.

